# Data quality and Big Data in the health industry: a scoping review protocol

**DOI:** 10.1101/2024.10.18.24315741

**Authors:** Lívia C. T Santos, Frederico M. Bublitz

## Abstract

**Introduction:** Big Data is characterized by the large volume of data, the variety of types and formats, the speed with which they are generated, and the veracity and value that can be extracted from the data. However, the result obtained with this technology will depend on the quality of the information obtained from the data. Big Data has great potential in healthcare and can be used to advance diagnosis, treatment, and healthcare management. Health data is highly vulnerable due to its sensitive nature, as it contains personal and confidential information. If exposed or compromised, it could lead to privacy violations, inaccuracies, misuse, incorrect diagnoses, or misguided decision-making in patient care. It is important to prioritize confidentiality, adhere to regulatory compliance, and maintain data integrity; for that, it is essential to use efficient methods to obtain quality data and make them able to reach the proposed objective.

**Objective:** In this context, the scoping review protocol aims to identify and map existing strategies, methods, or models that improve the quality of medical and health data in Big Data environments. This review explores the methods to support the effective use of Big Data in healthcare while addressing the challenges to maintain data integrity and ensure safe decision-making.

**Methods and analysis:** This scoping review will be conducted based on the six-step process outlined in the framework proposed by Levac et al. in “Scoping Studies: Advancing the methodology” and will be reported following the PRISMA-ScR (Preferred Reporting Items for Systematic Reviews and Meta-Analyses Extension for Scoping Reviews) checklist. The research team will use Data Quality, Big Data, and Health terms to search for primary studies in the Scopus Document Search, IEEE Xplore Digital Library, and ACM Digital Library databases.

## INTRODUCTION

Data generation has significantly increased in today’s digital world due to the diverse methods available for acquiring, managing, and processing information [1]. This has led businesses and organizations to face the challenge of handling large amounts of data from various sources while extracting valuable insights from it [2]. Taurion [3] defines Big Data as an equation combining volume, variety, velocity, and veracity, with all these factors adding value. The concept of Big Data is characterized by the 5 V’s [4], which capture its essential features: (i) Volume refers to the large amount of data generated and obtained; (ii) Variety refers to the data types and formats available; (iii) Velocity refers to the rapid rate at which data is produced, transmitted, stored, and processed, emphasizing the need for real-time or near-real-time analysis; (iv) Veracity addresses the consistency and trustworthiness of the data; (v) Value refers to the insights and potential advantages derived from the data.

Big Data allows for more data-driven decision-making and innovation across numerous sectors. These innovations include using Large Language Models (LLMs) that use machine learning and deep learning to analyze vast amounts of text data and generate human-like language, detect patterns, and provide insights [5]. LLM can be applied to medicine, assisting with drafting medical documentation and diagnosis and improving communication between healthcare providers and patients [6].

In healthcare, Big data plays a critical role by supporting the analysis of large datasets from electronic health records, medical imaging, and wearable devices to help professionals make decisions and improve patient diagnosis and treatment. However, ensuring the reliability of health data remains a challenge, as the quality of these vast datasets directly impacts the effectiveness of big data-driven healthcare solutions in life-or-death decisions [7]. Some benefits of using this technology in healthcare include disease prediction [8,9] and enhancing personalized treatment for individual patients through Precision Medicine [10].

The size and speed of data generation also present challenges, particularly regarding data quality, integration, and privacy concerns. Cai and Zhu [11] highlight several challenges related to big data quality, including integrating diverse and complex data types that complicate the overall data management process; as the volume of data overgrows, evaluating its quality becomes increasingly challenging; furthermore, the fast-changing nature of data requires agile processing technologies to keep up with constant changes; another critical issue is the lack of unified standards for assessing data quality.

This article presents a scoping review protocol to address these challenges and improve the understanding of data quality and Big Data in the health industry. The protocol aims to systematically explore the existing literature on data quality and enhance data quality management strategies to promote the better use of big data in healthcare. This review will allow us to map the main problems or concerns with methods that mitigate those problems. Additionally, understanding the gaps in data quality research within the health industry is critical for guiding future research.

### Previous systematic reviews

During the search rounds to define the terms used in the search string, no systematic reviews were identified whose emphasis is understanding and listing the main models, frameworks, or criteria used in data quality for Big Data, whose focus is health systems.

Previous reviews have focused on identifying this technology’s benefits, issues, and possible applications in health [12-15].

## METHODS AND ANALYSIS

The scoping review will be developed following the methodological framework proposed by Levac et al. [16] in “Scoping studies: Advancing the methodology”, which provides recommendations to clarify and improve the scoping review methodology presented by Arksey and O’Malley [17]. The framework has six stages:

1. Identifying the research question.
2. Identifying relevant studies.
3. Study selection.
4. Charting the data.
5. Collating, summarising, and reporting the results.
6. Consultation (optional stage).

The PRISMA-ScR [18] checklist will also be used to report the results.

### 1. Identifying the research question

This research aims to determine criteria, processes, methods, or models applied to medical and health data quality for Big Data. From the proposed objective, this research seeks to answer the following questions:

- What are the main problems or concerns about the quality of Big Data in the health industry? As an initial step in the study of the use of big data in health, it is necessary to understand the difficulties encountered in using this technology in healthcare.
- What methods are used for data quality? This question aimed to identify the methods used in papers and their possible benefits.
- What problems with data quality does this method mitigate? Identifying the mitigated issues will allow us to categorize the challenges encountered using this technology.
- What gaps exist in the health industry’s data quality research for Big Data? Understanding the gaps will provide information for future work in this study area.

### 2. Identifying relevant studies

This review will be conducted in the Scopus Document Search^1^, IEEE XploreDigital Library^2^, and ACM Digital Library^3^ databases, which are multidisciplinary databases with extensive work related to this review’s scope.

The team will conduct a series of searches for primary studies relevant to the research with different terms associated with data quality, big data, and health.

As a result of rounds of search and evaluation of the relevance of the papers obtained, the final search string was defined using the terms ((“big data”) AND (“clean*” OR “quality” OR “dirty”) AND (“criteria” OR “process” OR “method” OR “model”) AND (“health*” OR “medic*”)) in databases.

### 3. Study selection

This review aims to identify different models, frameworks, or criteria for improving the quality of medical and health data in Big Data. The inclusion (INC) and exclusion (EXC) criteria in Table 1 will guide the search and review of the papers obtained and determine the study’s eligibility.

**Table 1:** Inclusion and Exclusion Criteria.

| Inclusion Criteria | Exclusion Criteria |
| --- | --- |
| INC.01 - Domain: The article must address criteria, processes, methods, or models applied to the quality of health data for Big Data.<br>INC.02 - Language: The article must be written in English.<br>INC.03 - Date: There are no date restrictions.<br>INC.04 - Study design: The article presents a primary study.<br>INC.05 - Availability: The complete article is available<sup>4</sup>. | EXC.01 - Duplicity: The same article is found in another database. In this scenario, duplicates will be removed.<br>EXC.02 - Does not meet all the inclusion criteria. |

With all the articles obtained from the searches in the databases and the eligibility criteria defined, the team will carry out the following steps:

1. Remove duplicate articles, which will be automatically identified after importing search results into Rayyan^5^, software for managing systematic reviews.
2. Reading the title and abstract and removing those studies that do not meet the inclusion criteria.
3. Perform a full-text review, excluding studies that do not meet the criteria and explaining the reasons for exclusion.

In case of conflict in the steps, the team member will discuss with the others to find a solution. The results obtained after following all the steps will complete the PRISMA flow diagram, which shows the number of articles identified, duplicated, screened, eligible, and included in the review.

### 4. Charting the data

After selecting the studies, data will be extracted and charted from the articles that meet the inclusion criteria. Two researchers will collect the information separately using a data-charting form and continually update it to expand as new important data is identified [16] and later merge it. The form will include:

- Article title
- Author(s)
- Country
- Year
- Publisher
- Study objectives
- Methodologies
- Results
- Data quality methods
- Characteristics of the techniques
- Data quality problems mitigated
- Study strengths and limitations
- Team conclusion about the article

The data-charting form will identify the main problems related to Big Data quality in the health industry by summarizing results and the specific quality issues addressed in each article. The methods used for data quality will be captured, allowing for a comparative analysis of their effectiveness and benefits. Additionally, by detailing the characteristics of these techniques and the problems mitigated, the review will categorize challenges encountered in utilizing Big Data in healthcare. Lastly, insights on study strengths and limitations will reveal gaps in current research, guiding future work in this area.

### 5. Collating, summarising and reporting the results

As defined in the methodological framework by Levac et al. [16], this stage contains three steps and will allow researchers to obtain meaningful information from the data collected.

- Analysis: The team will provide a qualitative and quantitative analysis of the extracted information. Qualitative can give a better overview of the techniques provided and open up new study opportunities. In the quantitative analysis, the classification of techniques will be carried out according to their characteristics and the mitigated problems;
- Reporting the results: The team will present the results using tables with findings and gaps;
- Findings: The aim is to discuss these results and possible applications in future research.

## DISCUSSION

This scoping review protocol will synthesize the existing literature and guide future reviews to answer the questions raised. It aims to identify the methods used to improve health and medical data quality for Big Data, determine the most used, and address the problems that mitigate them. The protocol will also map the principal concepts and gaps associated with the topic. The results obtained will help future work in this area of study.

## Data Availability

All data produced in the present work are contained in the manuscript

## AUTHOR CONTRIBUTIONS

All team members contributed equally to the conceptualization, development, and writing of the draft of the manuscript.

## ETHICS AND DISSEMINATION

No ethics approval is required for this scoping review protocol, as primary data will not be collected. The review will produce important outcomes in improving the quality of health data for Big Data. The results obtained from this scoping review will be published in a peer-reviewed journal.

## Footnotes

1 https://www.scopus.com/home.uri

2 https://ieeexplore.ieee.org/Xplore/home.jsp

3 https://dl.acm.org/

4 Articles must be available in the databases linked to the Portal of Journals of CAPES (Coordination for the Improvement of Higher Education Personnel).

5 https://www.rayyan.ai/

